# *Clostridium tetani* bacteraemia in the plague area in France: two cases

**DOI:** 10.1101/2024.02.09.24302441

**Authors:** M.A. Boualam, A. Bouri, M. Signoli, M. Drancourt, A. Caputo, E. Terrerzz, G. Aboudharam

**Affiliations:** Aix-Marseille University, IRD, MEPHI, AP-HM, IHU Méditerranée Infection, Marseille, France; IHU Méditerranée Infection, Marseille, France; Aix-Marseille University, CNRS, EFS, ADES, UMR, 7268 Marseille, France; Aix-Marseille University, École de Médecine Dentaire Marseille, France

**Keywords:** *Clostridium*, *Clostridium tetani*, tetanus, palaeomicrobiology, palaeoculturomics

## Abstract

**Background:** Paleoculturomics aims to culture ancient pathogens from human remains such as dental pulp, which traps a drop of blood at the time of the death, to diagnose bacteraemia. *Clostridium tetani* (*C. tetani*) bacteraemia is a rare situation, with only four case reports in the literature.

**Methods:** Fourteen teeth collected from 14 individuals buried at the site of the 1590 plague in Fedons, France, were surface decontaminated before the pulp was cultured under strict anaerobiosis with negative controls. Colonies were identified by mass spectrometry and whole genome sequencing, and *C. tetani-*specific PCR was performed using DNA extracted from dental pulps, calculus and sediments.

**Results:** *C. tetani* cultured in two dental pulp specimens from two individuals was firmly identified by MALDI-TOF mass spectrometry, and whole genome sequencing confirmed toxigenic *C. tetani*. In the remaining twelve individuals, no such *C. tetani* was recovered and further detection by PCR and palaeoculturomics of dental calculus and sediments surrounding the teeth in these two individuals remained negative.

**Conclusion:** Toxigenic *C. tetani* which did not result from mere environmental contamination, caused bacteraemia in two individuals from a modern time plague site in France.

## INTRODUCTION

*Clostridium tetani* (*C. tetani*) is a gram-positive, spore-forming, rod-shaped anaerobic bacterium the toxigenic strains of which cause tetanus (1). *C. tetani* forms spores with resistance to heat, desiccation and oxygen exposure, allowing *C. tetani* to survive for decades in environments such as soil (1). The *C. tetani* 2.8 Mb genome comprises a plasmid potentially encoding tetanus toxin TeNT (2)(3), and such toxigenic *C. tetani* strains cause deadly tetanus after local infection and, rarely, bacteraemia which has only been reported in four cases (**Table 1**).

**Table 1:**
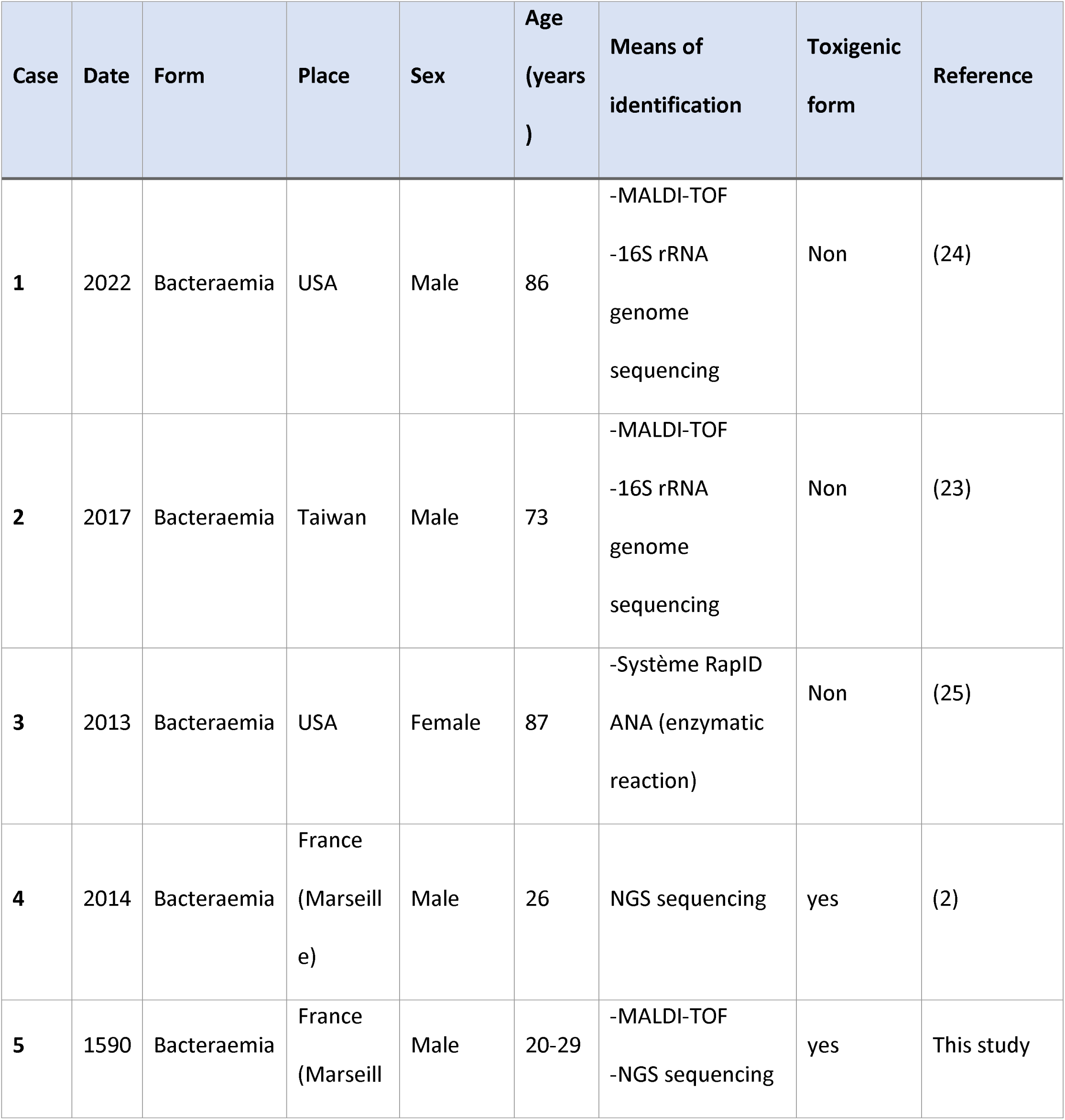
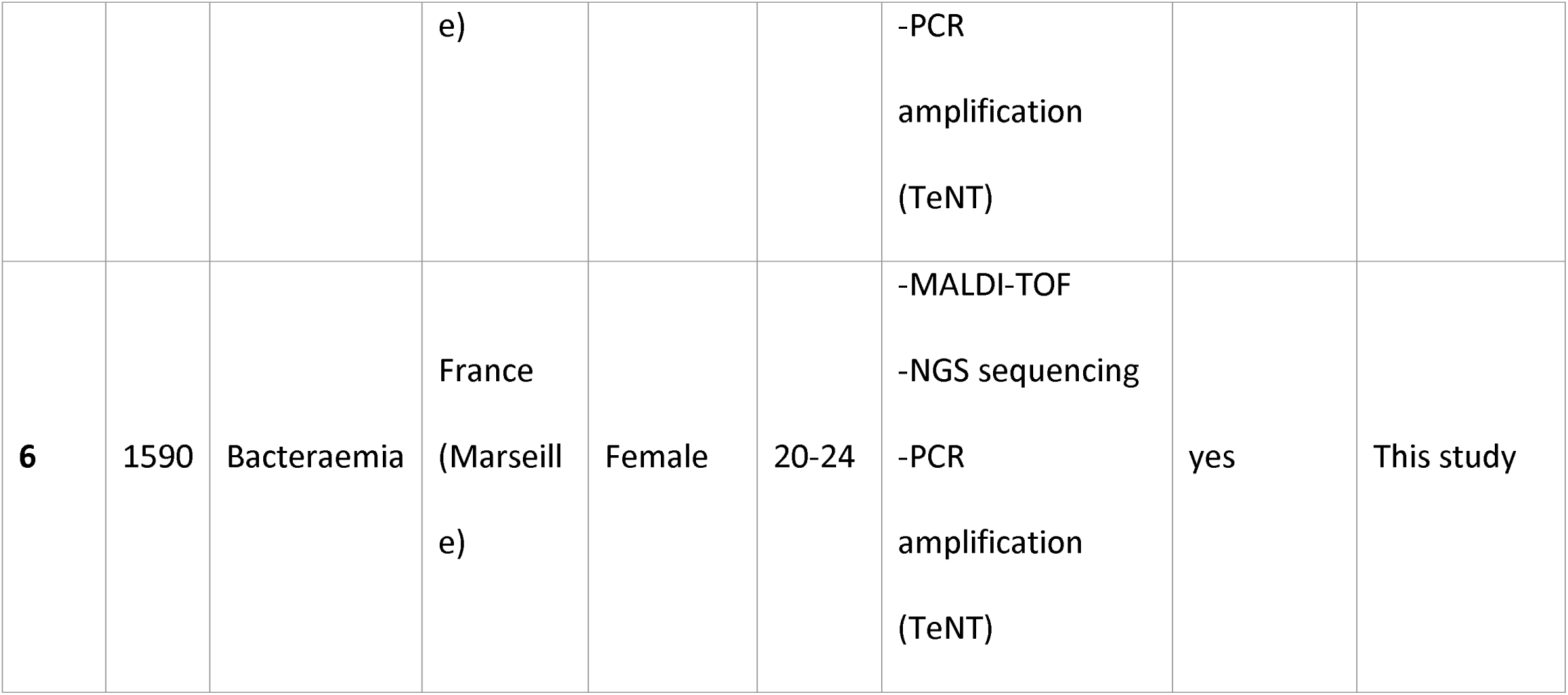
Reported invasive *Clostridium tetani* cases.

Here, our unanticipated observation of two additional cases of toxigenic *C. tetani* bacteraemia in two 16^th^ century individuals, questioned the natural history of this underreported form of *C. tetani* infection.

## MATERIALS AND METHODS

### Archaeological investigations

The Fédons burial site was discovered during preventive archaeology surveys carried out in anticipation of the construction of a new train line (4). This site is located 3.5 km west of the town of Lambesc in France (43°39′17″ North, 5°15′45″ East). The site is unambiguously related to a 1590 plague infirmary as deduced from confrontation of historical sources with archaeological and anthropological observations (5) and confirmed after palaeomicrobiological investigations firmly documented the causative *Yersinia pestis* (*Y. pestis*) by using aDNA investigations and immunochromatographic ones (5,6). This mass grave contained 133 skeletons, buried at the same time and in the same place (4). Fourteen tooth samples were collected from 14 of these individuals of different ages and sex, as determined using the methods presented by Schmitt A. *et al*., 2022 and Bruzek J. *et al*., 2002 (7, 8). For palaeomicrobiological investigations, one tooth from each individual was placed into an individual plastic bag with a note containing information on the individual and the sampling site. The teeth were kept at room temperature in a dedicated palaeomicrobiology laboratory at IHU Méditerranée Infection, Marseille, France, in accordance with French regulations for archaeological studies.

### Paleoculturomics

Each tooth yielded calculus before the external surface was disinfected with 99% ethanol and bleach for 30 seconds. All further steps were performed under an anaerobic hood (Don Whitley, Bingley, UK) in order to avoid exposure to atmospheric oxygen in the presence of negative controls, as previously described (9). All the instruments used for tooth opening and dental pulp extraction were sterilised before being placed under the anaerobic hood. Each tooth was cut in half, and the pulp of each half tooth was scraped into a 1.5 mL Eppendorf tube and hydrated with 10 µL of sterile phosphate buffered saline (PBS) (**Figure 1**). The same procedure was repeated with previously preserved dental calculus. Then, 5 µL of dental pulp and 5 µL of rehydrated dental calculus were separately inoculated onto a 5% sheep blood agar petri dish (Becton Dickinson GmbH, Heidelberg, Germany) and 5 µL onto Brain Heart Infusion Broth medium (Merck KGaA, Darmstadt, Germany) supplemented with haemin (Merck KGaA, Darmstadt, Germany) (**Figure 1**). The negative control consisted of 10 µL sterile PBS inoculated onto a 5% sheep blood agar plate. During the entire handling process, a culture medium plate was opened into the anaerobic hood to control for hood sterility. Each inoculated blood agar plate was placed in a bag with an anaerobic generator (BD GasPak, EZ Pouch Systems, Becton Dickinson, Franklin Lakes, NJ, US) and incubated at 37 °C under a 5% CO_2_ enriched atmosphere (3). Such bags also contained one negative control plate.

**Figure 1:**
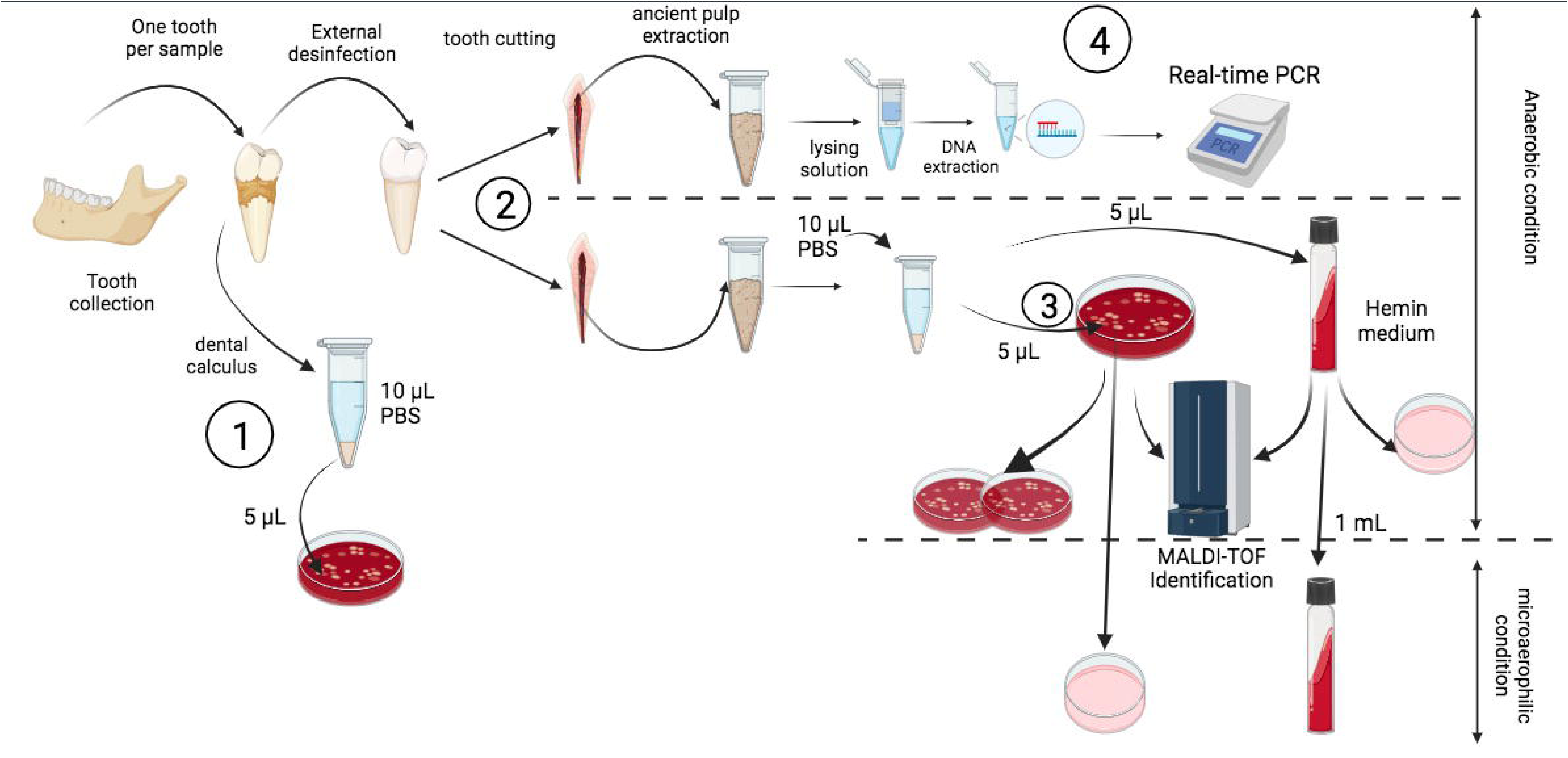
Workflow of the paleoculturomics approach, (**1**) cultivation of ancient dental calculus as soil contamination control. (**2**) Dental pulp extraction and pulp culture after hydration. (**3**) Identification of positive cultures. (**4**) DNA extraction and PCR screening.

### aDNA extraction

aDNA was extracted from dental pulp and calculus using a previous reported protocol, with modifications (10). In brief, 1 mL of lysis buffer (900 µL of 0.5M EDTA, 10 µL of 25 mg/mL proteinase K, 90 µL of nuclease free water) was incubated with pulp or dental calculus for 14 hours at 37 °C in a rotative wheel at 500 rpm. A negative control was performed from the beginning of extraction and consisted of nuclease free water. After incubation, centrifugation at 13 000 g for two minutes was performed and the supernatant was transferred to a sterile 50 mL Falcon tube containing10 mL of binding buffer (3.6 mL of nuclease free water, 7.16 g of guanidine hydrochloride powder, 6 mL of isopropanol, 150 µL of freshly prepared 5% Tween-20, and 450 µL of 3M sodium acetate (pH, 5.2) with vortex. Then, 700 µL of the solution was transferred on Qiagen MinElute Silica Spin columns, previously placed in a QIAvac vacuum systems (Qiagen, Hilden, Germany) after complete aspiration of the added solute and this step was repeated until the full 11 mL of the sample had been adsorbed on the column. To wash, we added 700 µL of PE buffer to the MinElute column and centrifuged for one minute at 13 000 g. To elute DNA, 12.5 µL of TE buffer were added with centrifugation for one minute at 13 000 g, this step was repeated twice in order to obtain a final volume of 25 µL.

### PCR-sequencing

To confirm the presence of *C. tetani* in the ancient pulps, a PCR targeting a 240-bp tetanospasmin (tent) gene of *C. tetani* was performed, using the following primers set by Guo *et al*, 2012 (11): F1-ATGCGCCATCGTATACTAAC; R1-CCATCTTTCGGATAACCTACA; F2-TATGTATTTGACAAATGCG; R2-CTTTCGGATAACCTACAAT. The thermal profile included an initial five-minute denaturation step at 95 °C followed by 45 cycles consisting of 30 seconds of dissociation at 95 °C, 30 seconds of annealing at 51 °C and 60 seconds elongation at 72 °C. Amplification products observed by electrophoresis gel migration were confirmed by sequencing after a purification step and sequence alignment against tetanus toxin and other *Clostridium* species toxins using MEGA X (12). The extraction negative control followed the same steps throughout the manipulation.

### Whole genome sequencing (WGS)

The two *C. tetani* isolates were further investigated by WGS using Illumina MiSeq (Illumina Inc., San Diego, CA, USA) following a previously reported protocol (13). Reads were trimmed by removing adapters using the CLC genomics workbench and decontaminated using BBduk tools from Galaxy Europe online software (Galaxy, https://usegalaxy.eu/) (14). The resulting reads were analysed online with taxonomic sequence classification Kraken2 software and visualised by Krona Pie chart on Galaxy Europe (**Supplementary data, Figure 1**). Generated reads were assembled using Unicycler software (15)and assembled genomes were blasted against the NCBI database to confirm identity. After obtaining the whole genome sequence, annotation was performed on the Bacterial and Viral Bioinformatics Resource Center (BV-BRC) (https://www.bv-brc.org) (16), including antibiotic resistance and virulence genes analysis (**Supplementary data, Figure 2**). Finally, taxonomic classification based on DNA-DNA hybridisation (DDH) was performed using Type (Strain) Genome Server (TYGS) (https://tygs.dsmz.de/) (17). After genome annotation using Prokka tools (18), genomic comparisons incorporating coding DNA sequences (CDS) extracted from 10 *C. tetani* genomic sequences [E88 reference (GCF_000007625.1), Havard (GCF_004119355.1), NIID-071400-001 (GCF_033128285.1), KHSU-254310-026 (GCF_033128265.1), KHSU-144316-041 (GCF_033128185.1), ATCC 453 (GCF_000762325.1), Mfbjulcb2 (GCF_003013635) and CMCC64008 (GCF_029636225.1) along with ancient genomes Q7452 (GCF_949357665.1) and Q7451 (GCF_949357675.1)] were made using an online protein sequence-based bidirectional BLAST approach on Proksee (https://proksee.ca/)(19). Resulting proteomes were analysed with Roary (Rapid large-scale prokaryotic pangenome Analysis) (20) to deduce pan-, core- and accessory genomes. Gene presence / absence and distribution of core and shell gene blocks were uploaded and visualised on Phandango (21) in the perspective of distinguishing any genomic differences between “ancient” and “modern” *C. tetani* genome sequences.

## RESULTS

Palaeoculturomic investigations of 14 dental pulps and 14 dental calculus yielded *C. tetani* in culture in dental pulps retrieved from two individuals, Ind-7 and Ind-44, whereas all the other cultures, including the negative controls, remained sterile after one week of incubation. Microscopic observation of colonies showed sporulated, gram-positive bacteria (**Figure 2**) identified as *C. tetani* by matrix-assisted laser desorption ionization time-of-flight mass spectrometry, with an identification score > 2.12 (22). Isolated *C. tetani* from Ind-7 and Ind-44 were deposited in the Collection de Souches de l’Unité des Rickettsies (IHU Méditerranée Infection, Marseille, France) under reference numbers CSUR Q7451 and CSUR Q7452, respectively. Further, toxigenic *C. tetani* strains were confirmed by aDNA PCR amplification and sequencing of a 240-bp TeNT fragment in Ind-7 and Ind-44 dental pulps (data not shown)(11). *C. tetani* was not detected by the two different paleoculturomics and paleomicrobiology approaches in the sediments and calculus surrounding the *C. tetani*-positive teeth. WGS yielded a 2 875 936-base pair (bp) genome with a 28.5% GC content for *C. tetani* CSUR Q7451 isolated from Ind-7 (accession number: GCA_949357675), and a 2 857 805-pb genome with a 28.6% GC content for *C. tetani* CSUR Q7452 isolated from Ind-44 (accession number: GCA_949357665). Genomic analysis confirmed *C. tetani* identification with an 87.9% DDH value with reference genome (GCF_000762305.1) for ancient isolates, both encoding a 3,948-bp, 27% GC content-tetanus neurotoxin *tet*X gene exhibiting 99.34% sequence similarity with homologous reference gene. Genomic comparisons revealed 2,268 core genes (present in 100% of studied genomes) forming 55.28% of the 4,103-gene pangenome, 1,094 shell genes (present in 15-95% of genomes) forming 26.67% and 741 cloud genes (present in < 15% of genomes) forming 18.05% of pangenome (**Figure 4**). Ancient Q7451 and Q7452 genomes specifically lacked 283 genes (**Figure 3**); and specifically incorporated three genetic blocks for a total of 4,780 bp (**Supplementary data Figure 3**; **Supplementary Table 1**). According to the anthropological description and the bone maturation, Ind-44 was a woman aged between 20 and 24 years at the time of her death and Ind-7 was a man aged between 20 and 29 years old at the time of his death.

**Figure 2:**
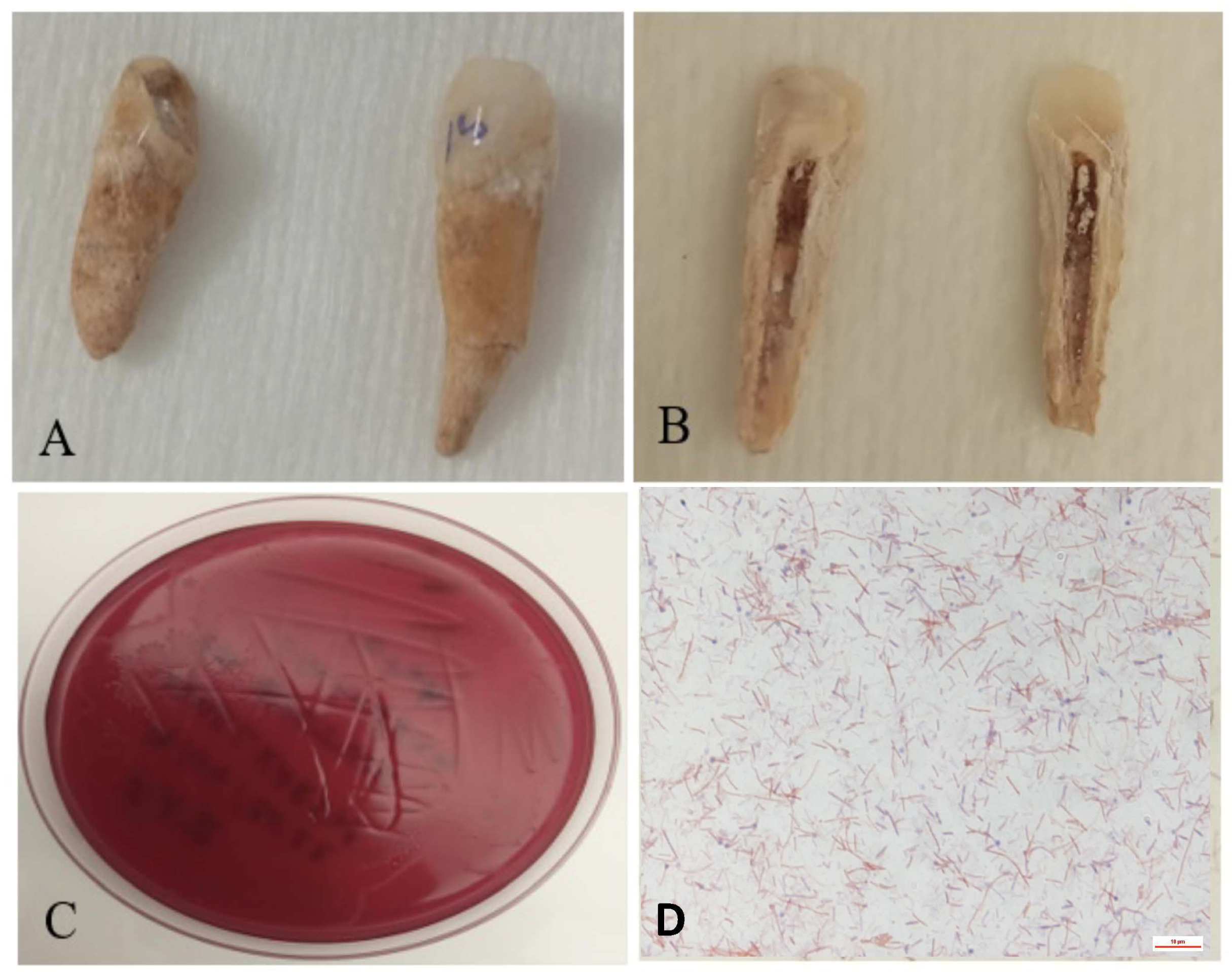
(**A-B**) Dental pulp sample from two immature individuals dated to the 16^th^ century. (**C-D**) Isolation bacteraemia of *Clostridium tetani*. (**A**) The 16^th^ century teeth of two prepubescent individuals are cleaned with 99% ethanol and bleach. (**B**) The dental pulp is kept in a closed environment (the pulp cavity). (**C**) Appearance of a bacterial carpet of *C. tetani* after one week of incubation on COS Agar enriched with 5% sheep blood. (**D**) Microscopic observation of *C. tetani* in rod form after staining with magnification 100X.

**Figure 3:**
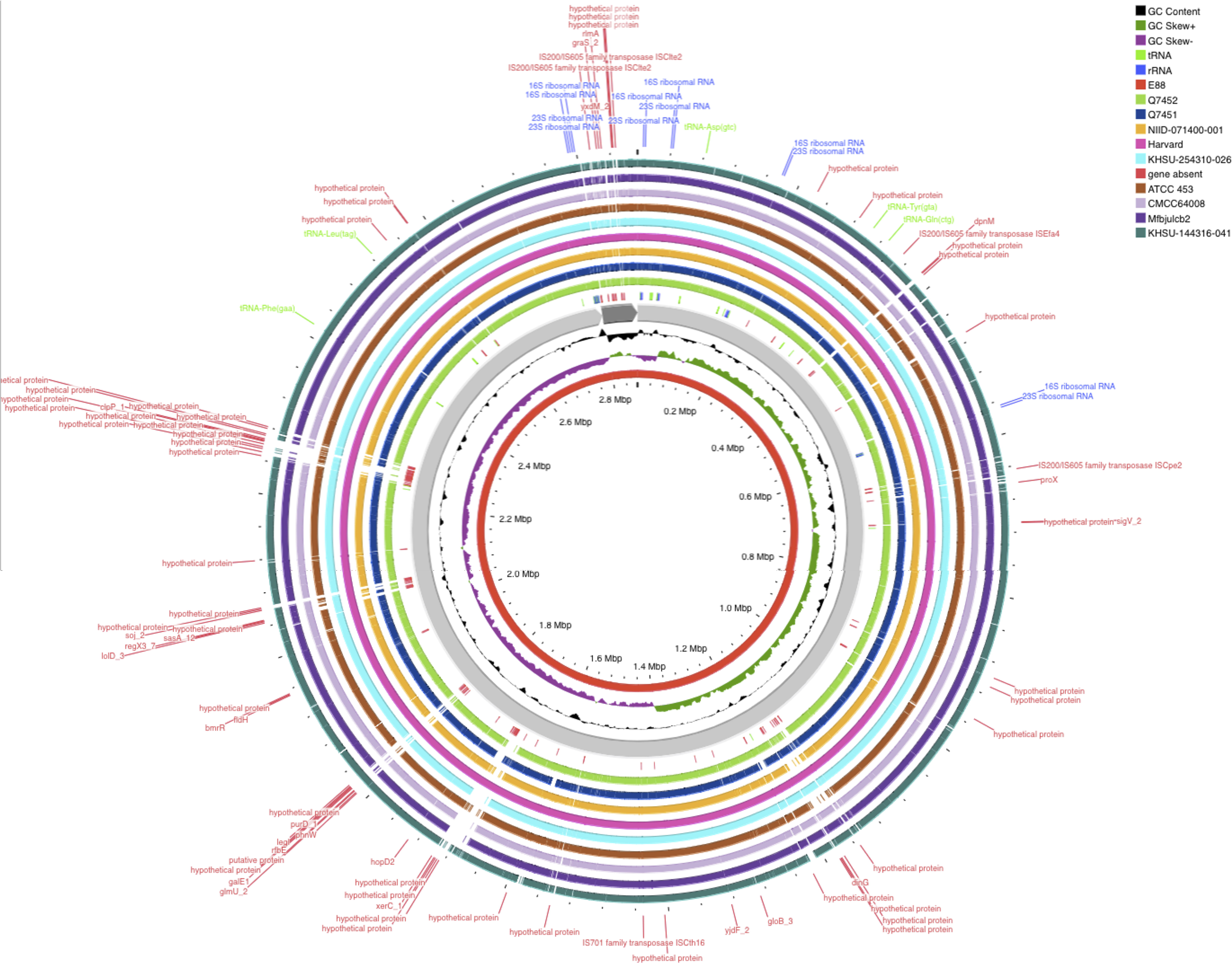
Circular representation of the genomic comparisons using coding DNA sequences (CDS), display on Proksee online (https://proksee.ca/)(19), displaying Blast comparison of the ten *C. tetani* strain genomes, features an in the in red ring in middle depicting the CDS of *C. tetani* reference strain E88, surrounded by two rings depict GC content and the GC skew and a gray ring representing the backbone. Following the backbone, the CDS present at the level of the reference genome and absent within the two ancient genomes Q7451 and Q7452 marked in red, tracking of the genes of the nine genomes including the two genomes reported in this study with different colors detailed in the legend.

**Figure 4:**
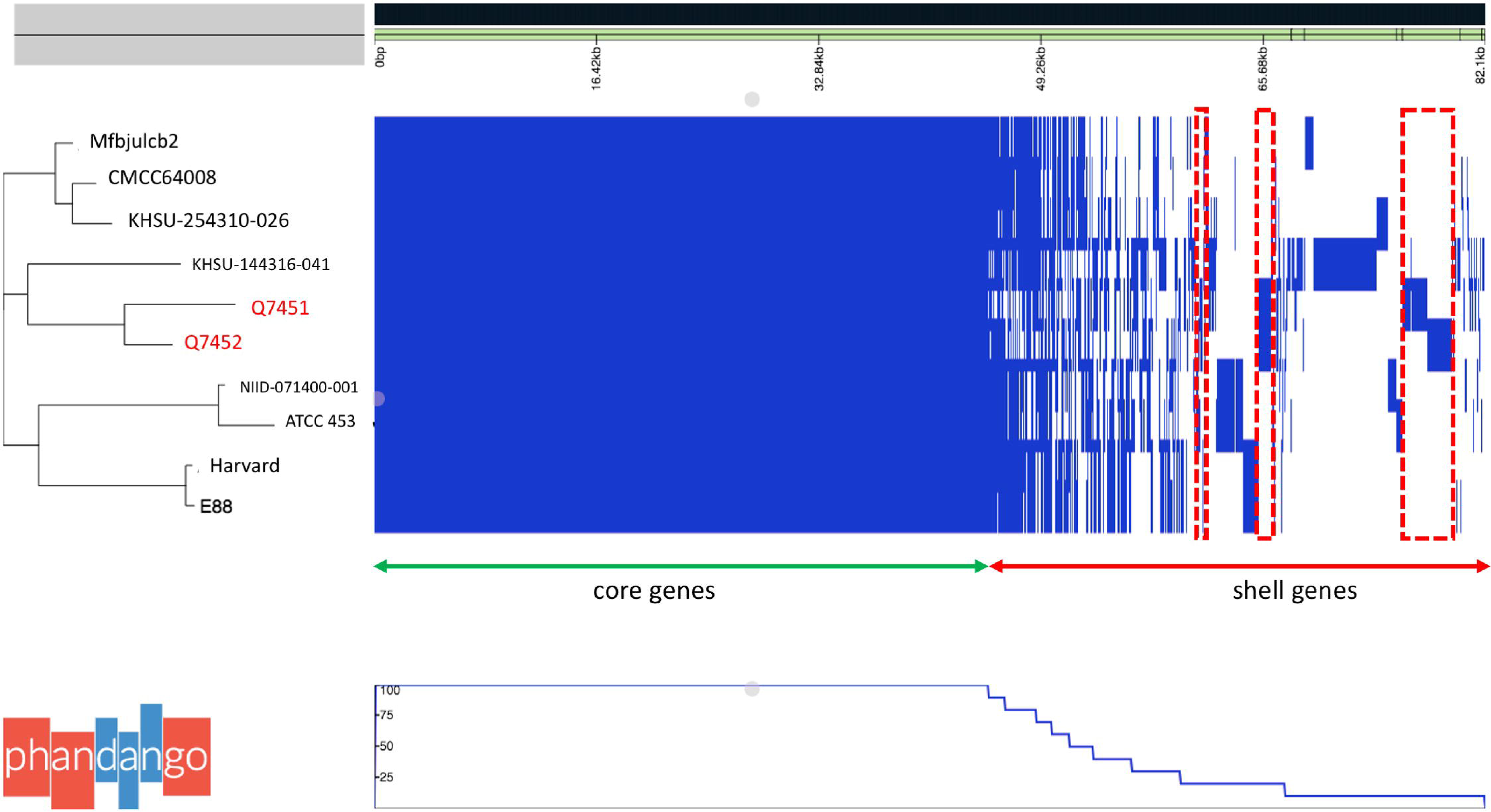
Pan-genome genetic relatedness analysis (utilizing the Roary pan-genome pipeline) (20,21), of the ten *C. tetani* strain. A total of 4,103 genes were identified. At left a dendrogram, in red the two strains of this study. Right: heatmap of the core genes each row shows the gene profile of each strain’s gene presence are in blue and gene absence in white matrix.

## DISCUSSION

In this study, culturing dental pulp collected from individuals who died during a 1590 plague episode in France (5, 6), resulted in the diagnosis of two cases of *C. tetani* bacteraemia. The non-detection of *C. tetani* (which had never been previously worked on in this laboratory), in surrounding soil and dental calculus specimens rendered external contamination of the dental pulp improbable, leading to the conclusion that bloodborne *C. tetani* contaminated the dental pulp at the time of death. Moreover, WGS comparisons of the two ancient genomes here reported with eight modern ones, indicated two lines of discriminant genomic events, common to the two ancient genomes and absent from modern counterparts, comforting the antiquity of strains Q7452 and Q7451. While deletions in ancient Q7452 and Q7451 genomes may testify of mis sequencing, the unanticipated discovery of three “in-block” DNA additions representing 12.2% of modern genomes clearly indicated that ancient Q7452 and Q7451 genomes genomically differed from modern ones and did not result from any mere contamination during the 500-year taphonomy process.

Indeed, highly vascularised dental pulp supported culturing a blood drop trapped inside the pulp cavity at the time of death. A sporulated form of *C. tetani* allowed the pathogen to survive for five centuries, a situation antedating by four centuries that of *Clostridium tertium* in 1914 soldiers ‘dental pulp in a quite a different historical and archaeological context (9). With the presence of appropriate controls, this observation of *C. tetani* recovered from two ancient pulp samples therefore led to the diagnosis of *C. tetani* bacteraemia, a condition rarely reported in the literature. Only four previous cases of such bacteraemia have been described in recent decades (**Table 1**). One case of *C. tetani* bacteraemia was in a 73-year-old man suffering from a hepatocellular carcinoma in 2017 in Taiwan (23). Another case was reported in an 86-year-old man with iatrogenic hypothyroidism in 2022 in the United States (24), and a third case was identified in an 87-year-old woman in 2013, also in the United States (25). In this study, toxigenic *C. tetani* bacteraemia was found in young individuals aged between 20 and 29 years at the time of their death, as calculated by bone maturity. A fourteen percent (2/14 individuals) prevalence of toxigenic *C. tetani* bacteraemia estimated in the present study, although derived from a small number of investigated cases, nevertheless may indicate a particular natural history of the pathogen in the context of plague.

Indeed, in this situation, Ind-7 but not Ind-44 tested positive for the plague agent *Y. pestis*, in line with previously reported detection of *Y. pestis* aDNA in this site (5). Present report is therefore one more illustration of dual infection in the course of documented plague, after cases of co-infection with *Y. pestis* has been reported with *Treponema pallidum* complex in a post-medieval and 17^th^ individual (26,27); with *Haemophilus influenzae* serotype b in 540 to 550 CE individual (28); with *Bartonella quintana* in individuals buried in a 11th–15th site in France (29). Furthermore, nine cases of *Streptococcus* spp.-*Y. pestis* coinfection have been reported between 1937 and 2019 (30). These studies question role of immunosuppressive effect of plague providing the opportunity for other pathogen replication, modulating their natural history. Present observation suggests that, in the context of plague, a deadly infection acutely diverting the immune system (31), *C. tetani* may act as an invasive agent, by-passing its portal of entry, an invasiveness attenuated in the contemporary population, and both pathogens contribute towards death. In these individuals, it is not possible to conclusively demonstrate route of infection. *C. tetani* spores are known to infect wound or injury with tetanus appearing several days later in case of toxicogenic strains (25). Most likely both individuals have contracted wound *C. tetani* infection before the death, with bacteriemia contributing to death. Indeed, surrounding sediments were negative for *C. tetani* following the palaeomicrobiological investigations, as reported.

This situation of dual infection may be underdiagnosed after detection of deadly plague may stop any further investigation for additional, potentially life-threatening pathogens, leading to neglect co-infection. Open-mind metagenomics and paleoproteomics investigations of ancient specimens as well as multiplex detection of pathogens may overcome misdiagnosis.

## CONCLUSIONS

Here, we diagnosed two cases of bacteraemia *C. tetani*, suggested a natural history of *C. tetani* infection in this population during the post-Black Death period, different from the natural history observed after the introduction of antitetanic serotherapy and vaccination in 1890 and 1924, respectively (32,33). In these anthropological and historical circumstances, *C. tetani* was probably an invasive agent with attenuated invasiveness in the contemporary population. The possibility of cultivating bacteria from archaeological dental material and from ancient dental pulp in particular opens up new perspectives for palaeomicrobiology. Bacteraemia-causing microorganisms, otherwise undetectable, can be highlighted or isolated by this method (9).

## Data Availability

All data produced in the present work are contained in the manuscript

