## Supplementary figures 1-2 for "*Clostridium tetani* bacteraemia in the plague area in France: two cases"

**Supplementary figure 1:** Krona Pie chart representing the taxonomic sequence classification by Kraken2 on galaxy Europe (Galaxy, https://usegalaxy.eu/)

I7 – Run Q7451

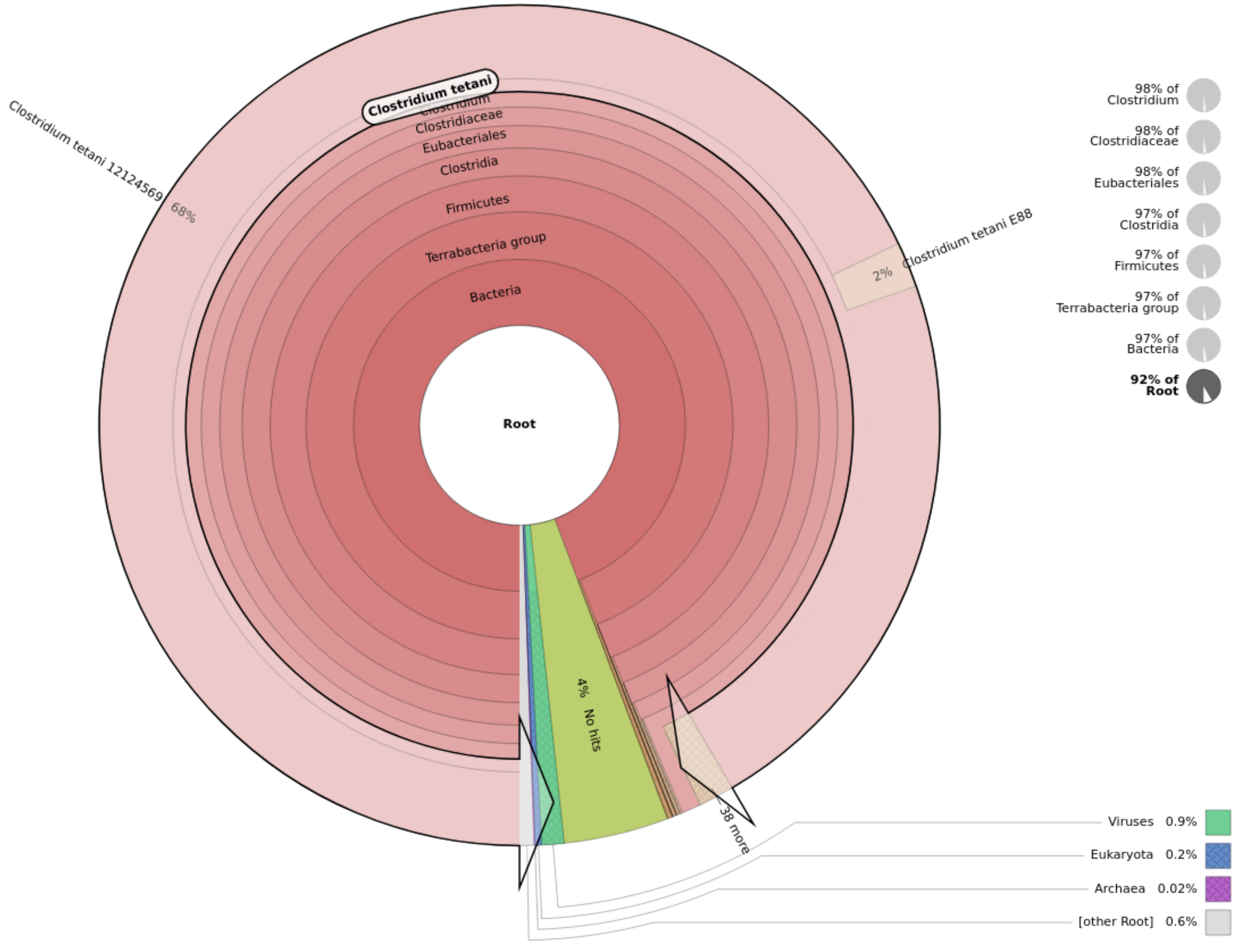

I44- Run 7452b

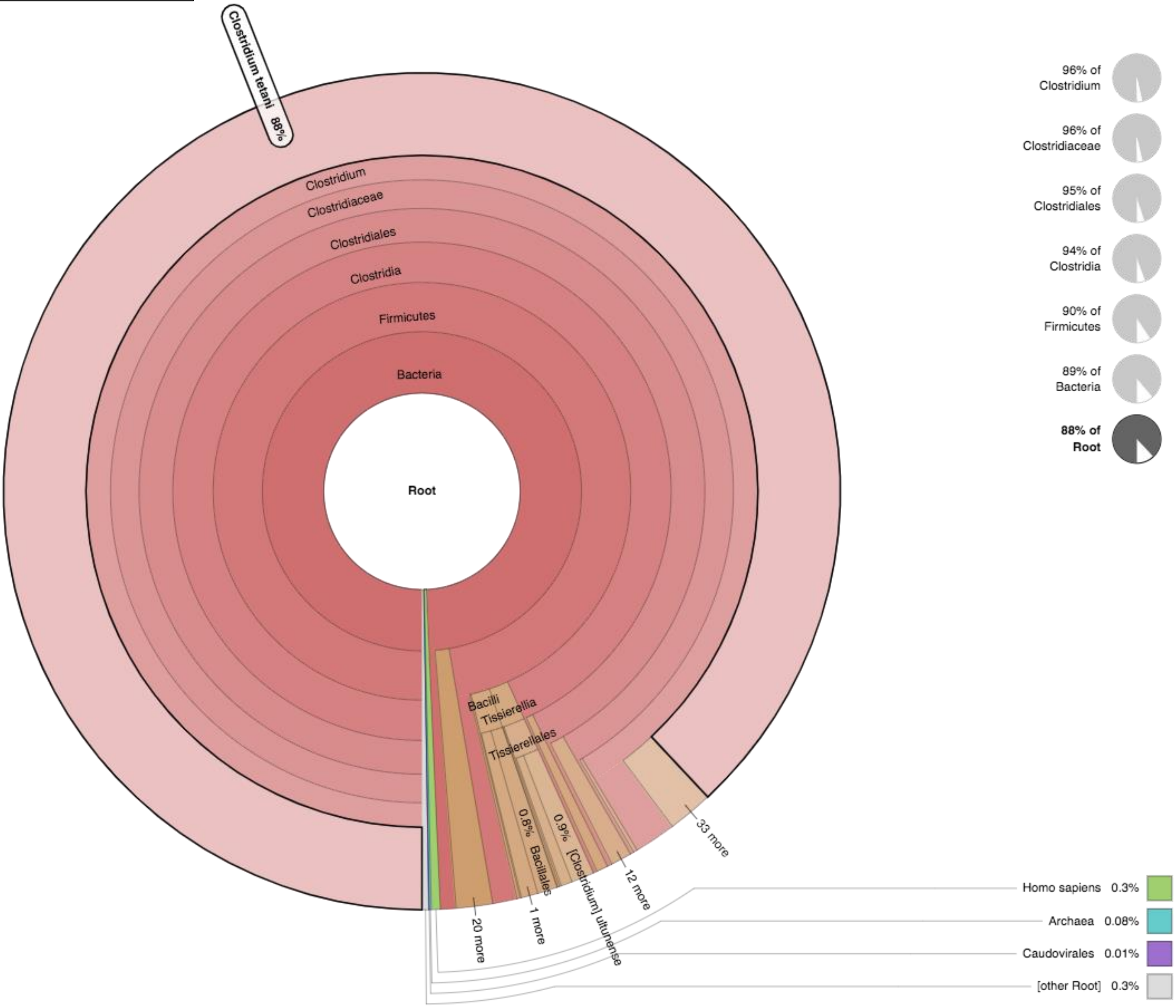

**Supplementary figure 2**:  generated whole genome Circular Viewer on the Bacterial and Viral Bioinformatics Resource Center (BV-BRC).

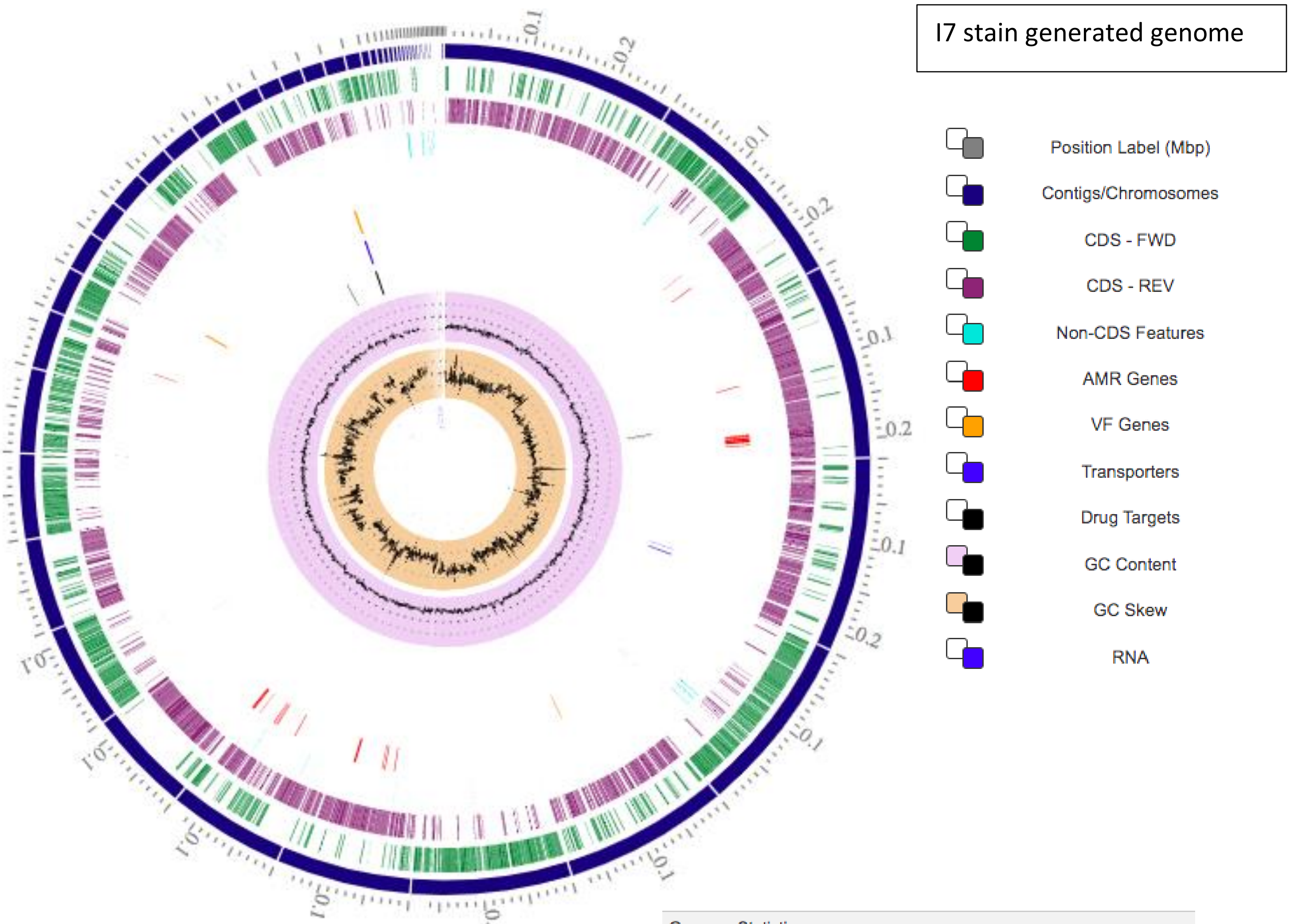


| | |
|---|---|
| Genome Statistics | |
| **Contigs** | 51 |
| **Genome Length** | 2880792 |
| **GC Content** | 28.504244 |
| **Contig L50** | 7 |
| **Contig N50** | 181715 |
| Annotation Statistics | |
| **tRNA** | 54 |
| **rRNA** | 7 |
| **CDS** | 2904 |
| **CDS Ratio** | 1.0080562 |
| **Hypothetical CDS** | 734 |
| **Hypothetical CDS Ratio** | 0.35640496 |
| **PLFAM CDS** | 2793 |
| **PLFAM CDS Ratio** | 0.96177685 |
| Genome Quality | |
| **Coarse Consistency** | 99.5 |
| **Fine Consistency** | 99 |
| **CheckM Completeness** | 99.8 |
| **CheckM Contamination** | 0.2 |
| **Genome Quality** | Good |
| Isolate Info | |

I44 stain generated genome

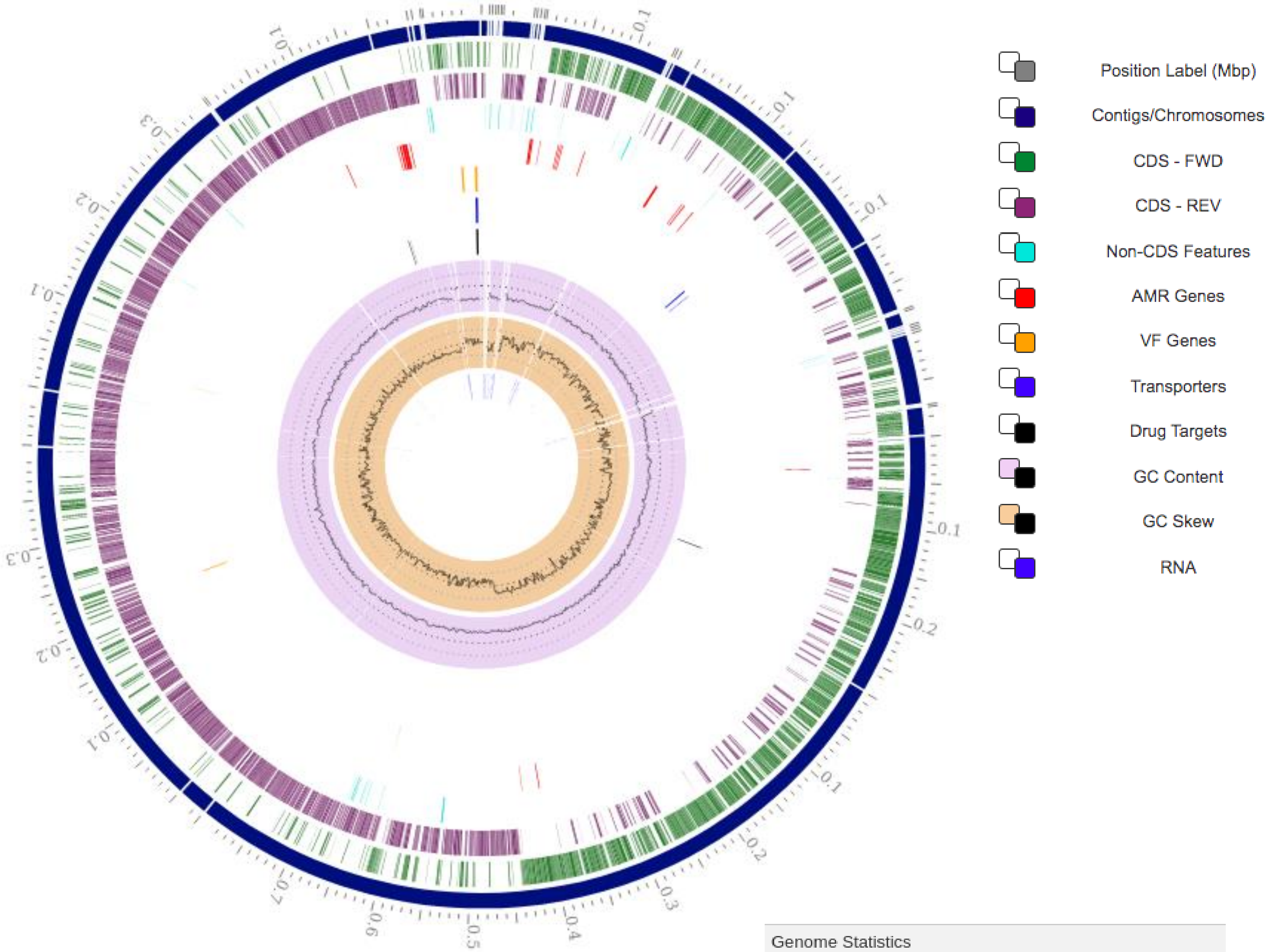


| | |
|---|---|
| Genome Statistics | |
| **Contigs** | 39 |
| **Genome Length** | 2858093 |
| **GC Content** | 28.68655 |
| **Contig L50** | 3 |
| **Contig N50** | 349193 |
| Annotation Statistics | |
| **tRNA** | 52 |
| **rRNA** | 14 |
| **CDS** | 2841 |
| **CDS Ratio** | 0.99401945 |
| **Hypothetical CDS** | 685 |
| **Hypothetical CDS Ratio** | 0.34494895 |
| **PLFAM CDS** | 2796 |
| **PLFAM CDS Ratio** | 0.9841605 |
| Genome Quality | |
| **Coarse Consistency** | 99.5 |
| **Fine Consistency** | 99 |
| **CheckM Completeness** | 100 |
| **CheckM Contamination** | 0.3 |
| **Genome Quality** | Good |
