## Supplemental Figure 3 for "*Clostridium tetani* bacteraemia in the plague area in France: two cases"

***Supplementary figure 3***

**Figure 3:** genomic comparisons using coding DNA sequences (CDS) by maps with multiple BLAST comparisons using Proksee online ([https://proksee.ca/](https://proksee.ca/)) (Grant et al., 2023), including height published *C. tetani* strains genomes : E88 (GCA_000007625.1) like reference genome, Havard (GCF_004119355.1), NIID-071400-001 (GCF_033128285.1), KHSU-254310-026 (GCF_033128265.1), KHSU-144316-041 (GCF_033128185.1), ATCC 453 (GCF_000762325.1), Mfbjulcb2 (GCF_003013635) and the two ancients *C. tetani* strains Q7452 (GCF_949357665.1) and Q7451 (GCF_949357675.1). (A) : whole genome comparison, (B) plasmid comparison, (C1-3) different very heterogeneous genetic islands between all strains showing genes absence in comparison with reference genome C. tetani E88.

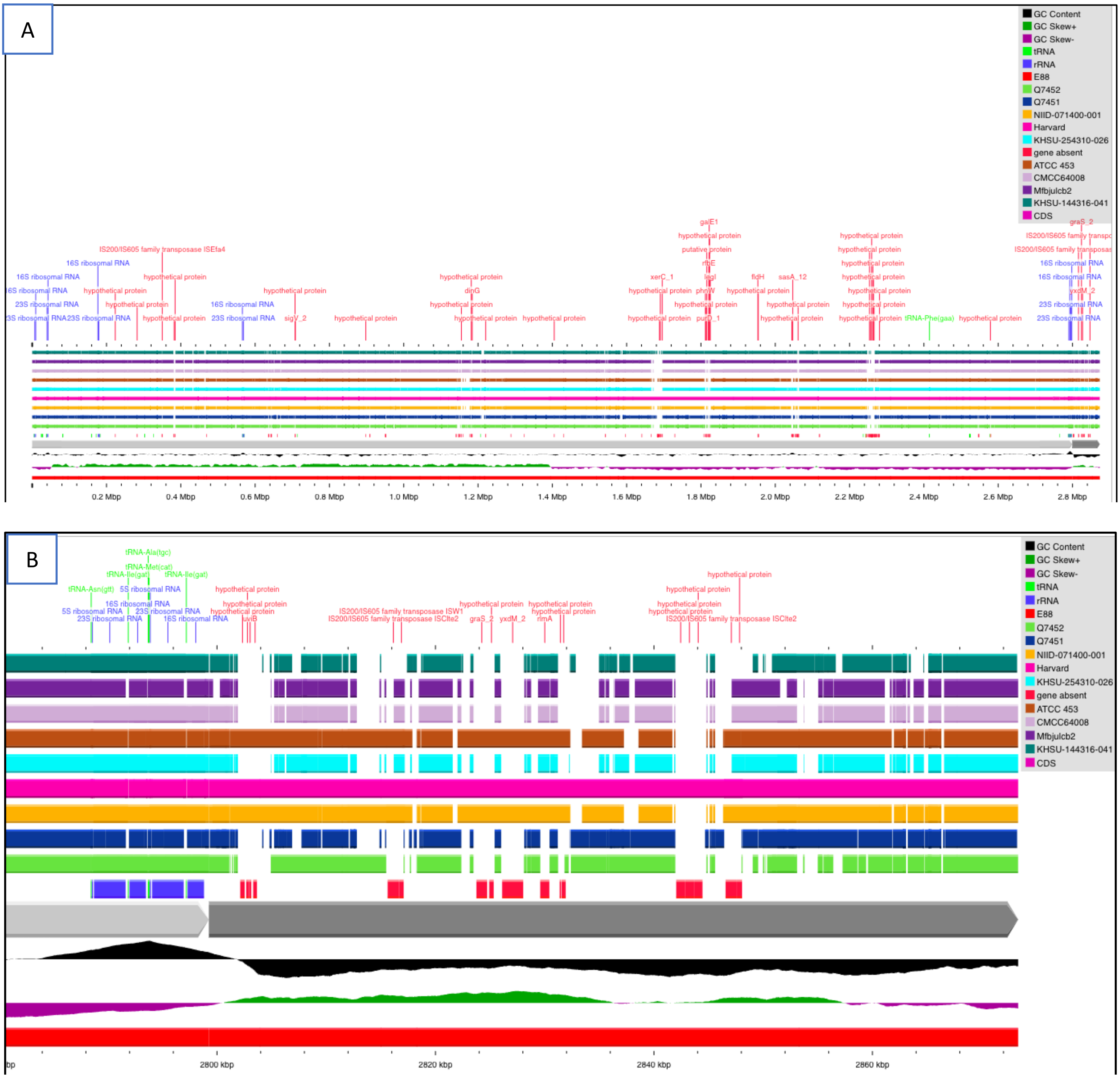

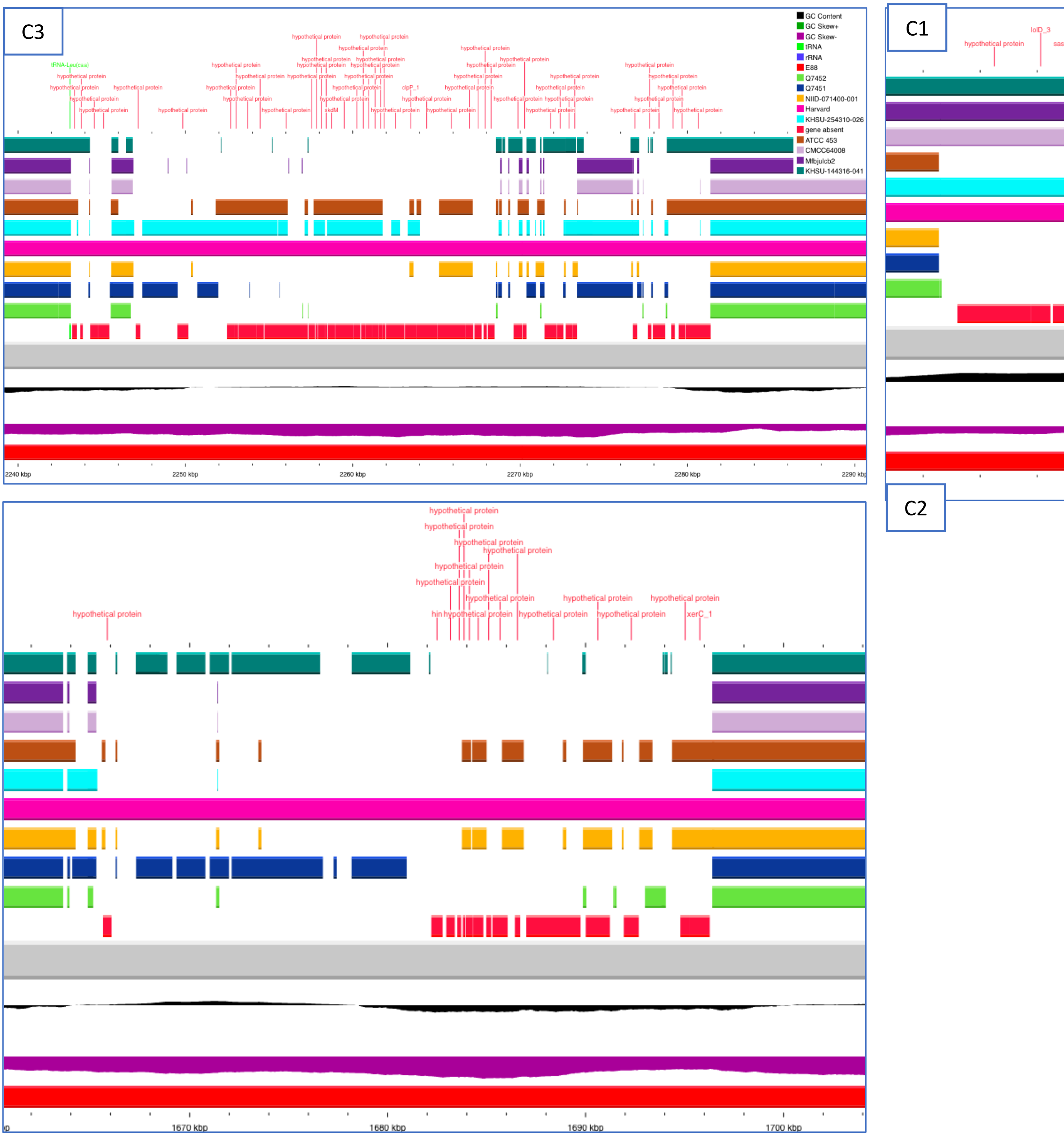
C3
GC Content
GC Skew+
GC Skew-
tRNA
rRNA
E88
Q7452
Q7451
NIID-071400-001
Harvard
KHSU-254310-026
gene absent
ATCC 453
CMCC64008
Mfbjulcb2
KHSU-144316-041
tRNA-Leu(caa)
hypothetical protein
clpP_1
xkdM
2240 kbp
2250 kbp
2260 kbp
2270 kbp
2280 kbp
C1
lolD_3
C2
xerC_1
1670 kbp
1680 kbp
1690 kbp
1700 kbp
